# Association of alcohol consumption with circulating biomarkers of atrial fibrillation-related pathways in a population at high cardiometabolic risk

**DOI:** 10.1101/2023.12.05.23299449

**Authors:** Aniqa B. Alam, Estefania Toledo-Atucha, Dora Romaguera, Angel M. Alonso-Gómez, Miguel A. Martínez-Gonzalez, Lucas Tojal-Sierra, Cristina Razquin, Marta Noris Mora, Linzi Li, Vinita Subramanya, Jordi Salas-Salvadó, Montserrat Fitó, Alvaro Alonso

**Affiliations:** Department of Epidemiology, Rollins School of Public Health, Emory University, Atlanta, GA, USA; CIBER Consortium, M.P. Physiopathology of Obesity and Nutrition (CIBERObn), Carlos III Health Institute (ISCIII), Madrid, Spain; Navarra’s Health Research Institute (IdiSNA), Navarra Institute for Health Research, Pamplona, Spain; University of Navarra, Department of Preventive Medicine and Public Health, Pamplona, Spain; Health Research Institute of the Balearic Islands (IdISBa), Palma de Mallorca, Spain; Bioaraba Health Research Institute, Osakidetza Basque Health Service, Araba University Hospital, University of the Basque Country UPV/EHU, Vitoria-Gasteiz, Spain; Human Nutrition Unit, Department of Biochemistry and Biotechnology, Rovira i Virigili University, Reus, Spain; Human Nutrition Unit, Pere Virgili Health Research Institute (IISPV), Reus, Spain; Cardiovascular Risk and Nutrition Group, Hospital del Mar Medical Research Institute (IMIM), Barcelona, Spain; Department of Cardiology, Hospital Universitari Son Espases, Palma, Spain

## Abstract

**Background:** The effect of alcohol consumption on cardiovascular health, including atrial fibrillation risk, remains controversial. Evaluating the association of alcohol consumption with circulating atrial fibrillation-related biomarkers may help better understand the relevant mechanistic underpinnings.

**Methods:** We studied 523 participants from 3 sites for the PREDIMED-Plus study, a weight-loss randomized intervention trial in metabolically unhealthy adults. N-terminal pro-B-type natriuretic protein (NTproBNP), high sensitivity troponin-T (hsTnT), high-sensitivity C-reactive protein (hsCRP), 3-nitrotyrosine (3-NT), and procollagen type 1 carboxy-terminal propeptide (PICP) were measured in fasting serum samples at baseline and years 3 and 5 of follow-up. We calculated alcohol consumption in drinks/day (1 drink = 14 grams alcohol) with validated food frequency questionnaires at each visit. Using multiple linear regression and mixed models we estimated the association of alcohol consumption with log-transformed biomarkers at baseline and longitudinally adjusting for potential confounders.

**Results:** Among 523 participants (mean age: 65 years, 40% female), mean alcohol consumption was 1 drink/day. Cross-sectionally, alcohol consumption was not associated with cardiac biomarker concentrations. Longitudinally, compared to non-consumers, heavy drinkers (≥4 drinks/day) had smaller increases in hsTnT (β: -0.11, 95%CI: -0.20, -0.01)and PICP (β: -0.15, 95%CI: -0.30, 0.01) over the 5-year follow-up. In contrast, those who increased alcohol consumption over the 5-year period experienced greater increases in hsCRP (β: 0.42, 95%CI: 0.11, 0.73) compared to those whose drinking behavior stayed the same.

**Conclusion:** Alcohol consumption was associated with complex changes in circulating biomarkers, including comparatively lower fibrotic and myocardial damage, but higher levels of overall inflammation over time. These results underscore the need for further research to better understand the effects of alcohol on cardiovascular health.

## INTRODUCTION

Drinking culture is a staple in modern society. Over 90% of people in the US have consumed alcohol at some point in their life, and international estimates put it at 80% across multiple countries.^1^ Alcohol dependence and abuse are of particular concern, especially when considering cardiovascular health. The literature on alcohol consumption and cardiovascular health are extensive, yet mixed, with some studies showing a beneficial effect,^2^ while others suggest a causal role in heart disease.^3^ The link between alcohol and atrial fibrillation (AF) has been less explored.

Nearly 35% of AF patients report alcohol preceding AF events, making it the most common trigger of AF.^4^ Reducing alcohol intake has been associated with reductions in the number of AF episodes as well as time spent in AF.^5^ Inflammation of the myocardium,^6^ atrial remodeling,^7^ and impaired strain function^8^ have all been associated with drinking. These changes in cardiac structure and function contrast with other observations suggesting a beneficial effect of low-to-moderate alcohol consumption on coronary heart disease and other cardiovascular diseases.^3,9,10^ Thus, evaluating the association of alcohol consumption with selected biomarkers of AF-related pathways can help characterize the complex association of alcohol with cardiovascular disease in general, and AF in particular.

## METHODS

### Study population

The PREDIMED-Plus is an ongoing, multicenter, randomized control trial with the primary objective of assessing the effect of an intensive weight-loss intervention alongside an energy-reduced Mediterranean diet (erMedDiet) and behavioral intervention on the primary prevention of cardiovascular disease in 6,874 persons with metabolic syndrome.^11^ Those assigned to the control group were guided to follow the MedDiet without mention of energy reduction or physical activity. All Institutional Review Boards at participating institutions reviewed and approved this study’s protocol, and all participants have provided written informed consent to be part of the study.

This analysis was restricted to 538 participants from three study sites with available biomarker data. **Figure 1** provides a flowchart of selection into the current analysis. After excluding those who had AF at baseline (N = 12) and were missing other important demographic information (N = 3), 523 participants were included. Of these, 495 (95%) and 478 (91%) had biomarker data available at the year 3 and year 5 visits, respectively.

**Figure 1.**
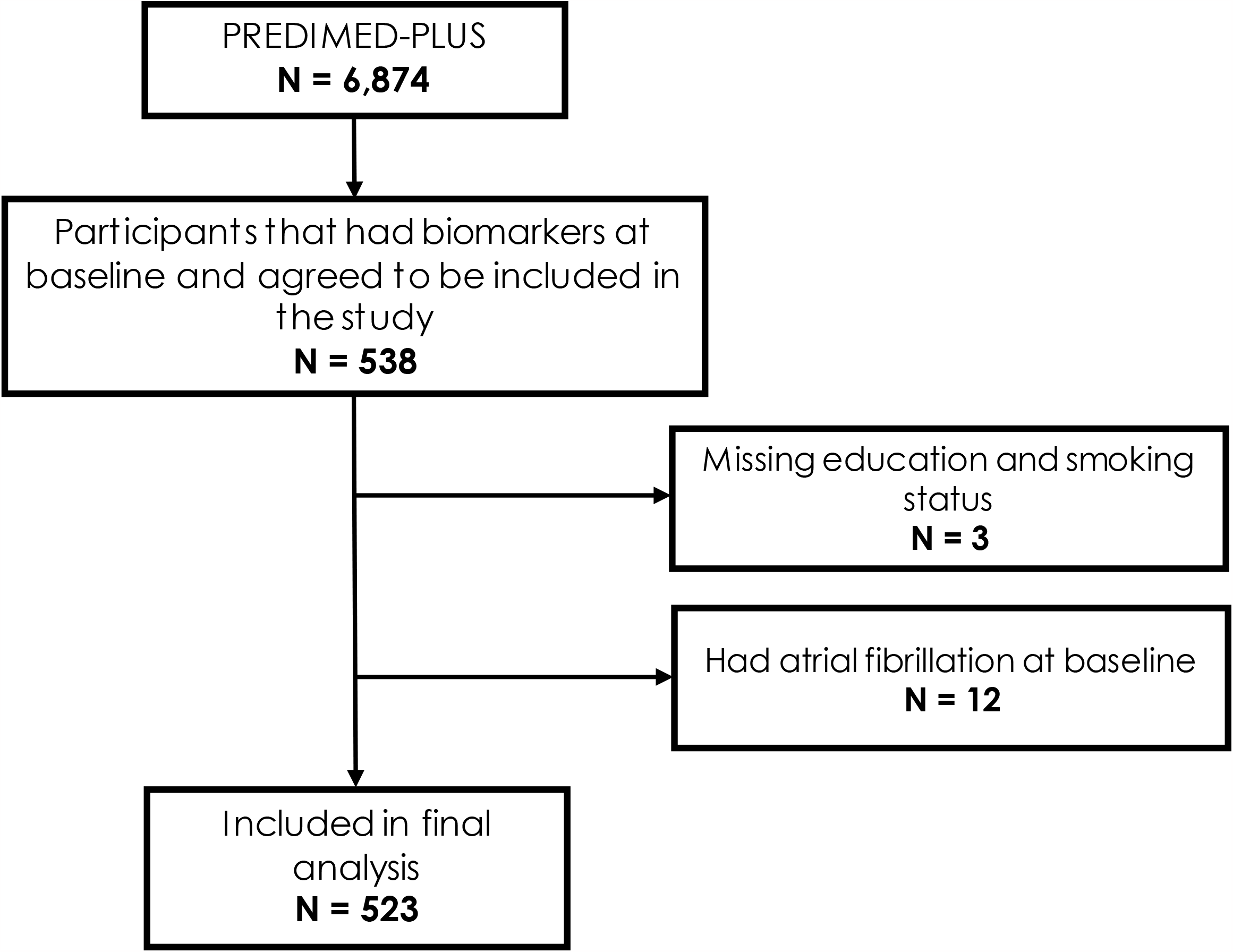
Flow chart of selection into the study.

### Cardiovascular Biomarkers

Biomarkers of interest for this analysis are purported to be involved in AF and AF-related pathways. These biomarkers include N-terminal pro-B-type natriuretic protein (NTproBNP) and high-sensitivity troponin-T (hsTnT), markers of myocardial stress and damage; high-sensitivity C-reactive protein (hsCRP), an indicator of inflammation; 3-nitrotyrosine (3-NT), a byproduct of nitrative stress and biomarker of oxidative stress; and procollagen type 1 carboxy-terminal propeptide (PICP), a marker of cardiac fibrosis. Biomarkers were measured in blood samples from fasting participants at baseline, year 3, and year 5. NTproBNP and hsTnT were measured by way of electrochemiluminescence immunoassay, while hsCRP was measured through immunoturbidimetry and were run on a Cobas 8000 autoanalyzer (Roche Diagnostics). Enzyme-linked immunosorbent assay technique (ELISA) was utilized to measure 3-NT (Human Nitrotyrosin ELISA kit; Abcam, Cambridge, UK) and PICP (MicroVue PICP EIA; Quidel, San Diego, CA, USA). The intra- and inter-assay variability for biomarkers are as follows: 3.8% and 6.9% for NTproBNP; 2.0% and 3.7% for hsTnT; 3.2% and 3.9% for hsCRP; 5.5% and 7.2% for PICP; and 10% and 15% for 3-NT.

### Alcohol Consumption

A validated 143-item food frequency questionnaire (FFQ) administered at every visit during the course of the trial was used to calculate usual alcohol consumption.^12^ One standard drink was defined as 14 grams of pure alcohol. As pure alcohol content is dependent on the type of alcohol, standard sizes for each type of beverage was defined as the following: 1 glass of wine (100 cc), save for muscatel (50 cc); 1 bottle of beer (330 cc); 1 shot of liquor or whiskey (50 cc). The intra-class correlation between FFQs and four 3-day dietary data was 0.82 for alcohol intake.^12^

### Covariates

All covariates are from baseline data collection. Education, marital status, and smoking status were self-reported. Metabolic equivalents of task (METs) in minutes per day served as measurement of physical activity.^13^ Body-mass index (BMI) was calculated from height and weight measured at baseline. Systolic and diastolic blood pressure were measured 3 times at each visit and then averaged. Diabetes status was determined through self-report, the use of antidiabetic medication, and measurements of fasting blood glucose and glycated hemoglobin.

The 21-question Beck Depression Inventory detected the presence of depressive symptoms. Finally, adherence to the MedDiet was assessed through a 16-item questionnaire; the original 17-item version of the questionnaire was edited to remove alcohol intake in order to avoid redundancy.^14^

### Statistical Analysis

We used multivariable linear regression models to estimate cross-sectional associations between alcohol consumption and cardiac biomarkers at baseline. We used mixed models to estimate longitudinal associations of baseline alcohol consumption with change in cardiac biomarkers from baseline to year 3 to year 5. All biomarkers were log-transformed and time was modeled in 5-year increments in longitudinal analyses. Alcohol consumption was categorized as no drinking (0 drinks/day, reference group), 1, 2-3, or ≥4 drinks/ day. Additional analyses were conducted with alcohol modeled as a continuous variable and with visit considered as a categorical variable (baseline, year 3, year 5). To assess the impact of change in drinking behavior over time, we used the difference in alcohol consumption from baseline to year 5 as the predictor of change in biomarkers over the course of the study. The difference in alcohol consumption was categorized into decreased consumption (≥ 1 drinks/day), no change (<1 drinks/day), and increased consumption (≥ 1 drinks/day).

Models were first adjusted for age, sex, education, and assignment to the intervention group. They were later also adjusted for marital status, smoking, physical activity, height, BMI, systolic and diastolic blood pressure, diabetes, depression, and adherence to the erMedDiet. Longitudinal analyses further accounted for interaction of all covariates with time. We performed additional analyses stratified by sex and tested for interaction including alcohol*sex terms in the models.

All analyses were performed using SAS 9.4 (Cary, NC; SAS Institute Inc.), while splines were generated using STATA, version 17 (StataCorp LP, College Station, Texas).

## RESULTS

Of the 523 participants included in this study, 40% were female and the mean age (SD) was 65 (5) years. Heavy drinkers were more likely to be male, more educated, and also more likely to have previously smoked or be currently smoking (**Table 1**). Red wine was the preferred drink of choice.

**Table 1.**
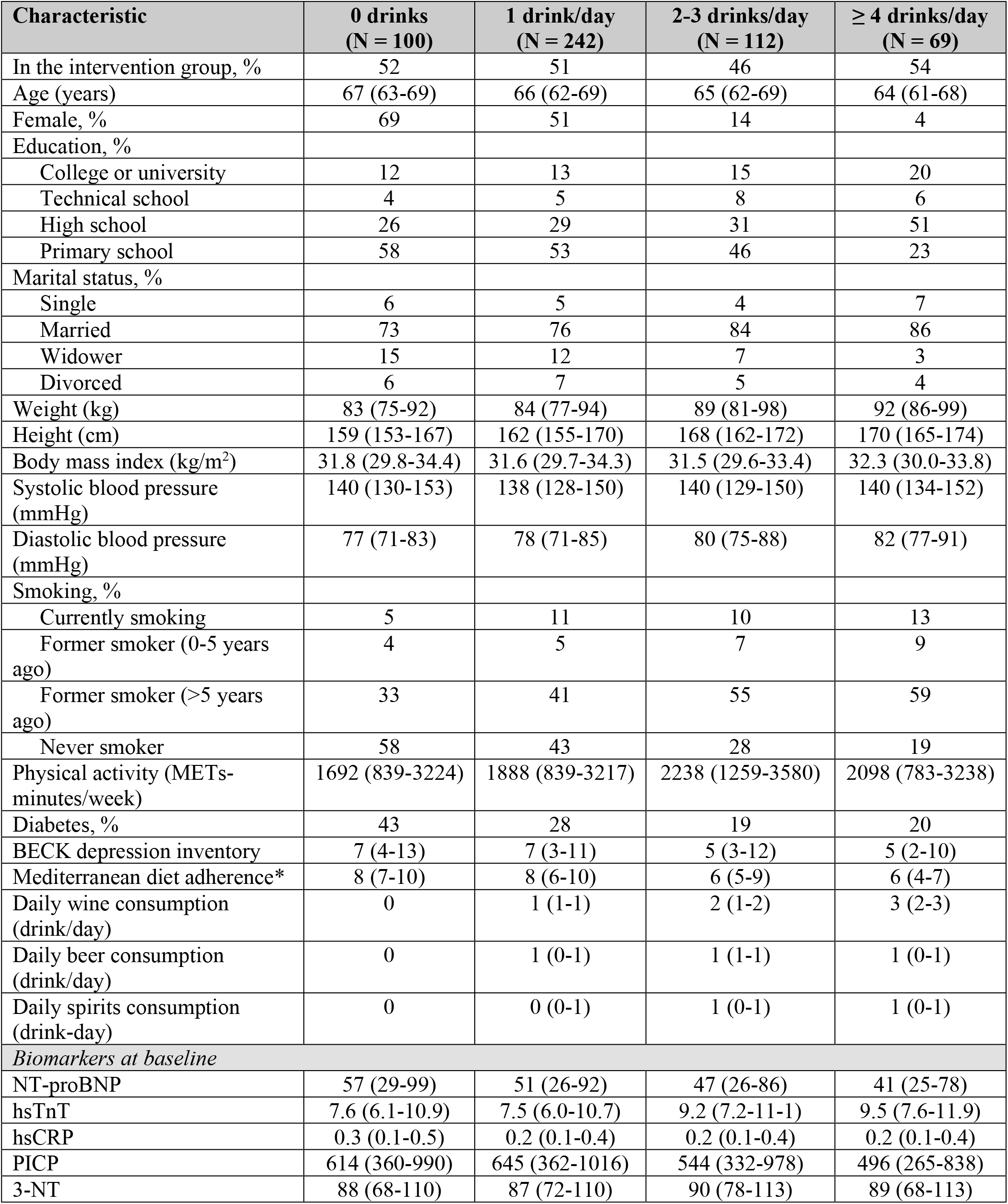
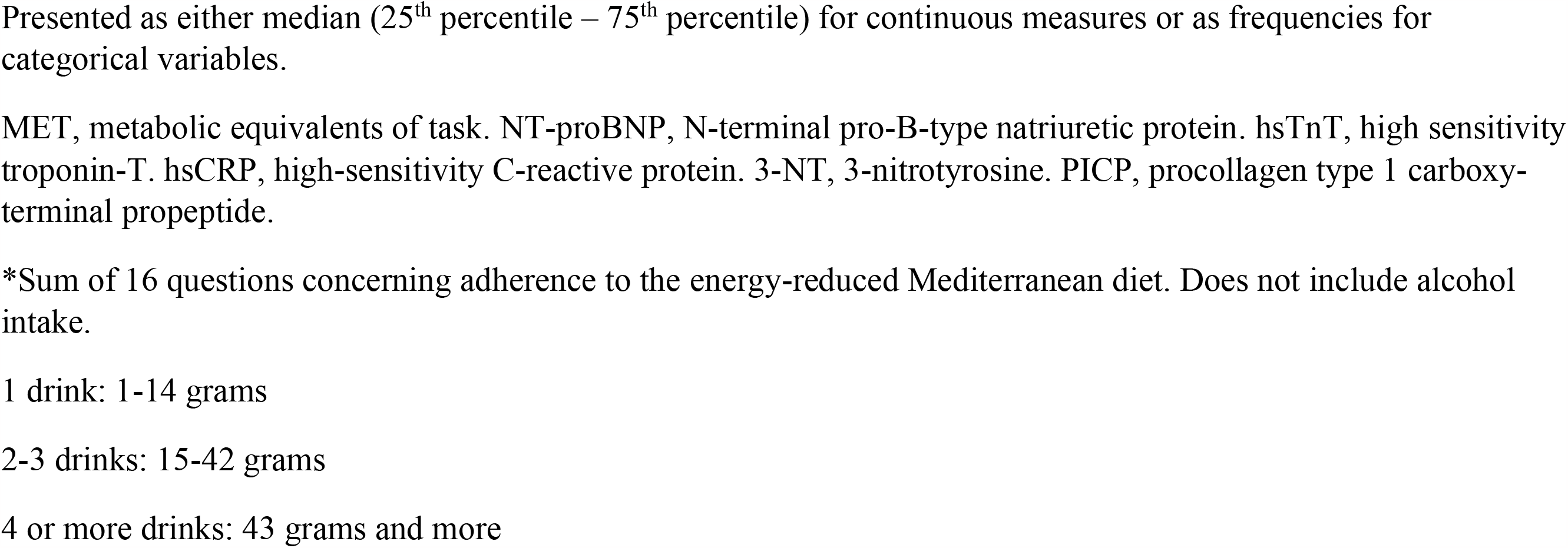
Baseline characteristics of study participants by baseline daily alcohol consumption (1 drink = 14 grams of pure alcohol), PREDIMED-Plus study.

| Characteristic | 0 drinks<br>(N = 100) | 1 drink/day<br>(N = 242) | 2-3 drinks/day<br>(N = 112) | ≥ 4 drinks/day<br>(N = 69) |
| --- | --- | --- | --- | --- |
| In the intervention group, % | 52 | 51 | 46 | 54 |
| Age (years) | 67 (63-69) | 66 (62-69) | 65 (62-69) | 64 (61-68) |
| Female, % | 69 | 51 | 14 | 4 |
| Education, % |  |  |  |  |
| College or university | 12 | 13 | 15 | 20 |
| Technical school | 4 | 5 | 8 | 6 |
| High school | 26 | 29 | 31 | 51 |
| Primary school | 58 | 53 | 46 | 23 |
| Marital status, % |  |  |  |  |
| Single | 6 | 5 | 4 | 7 |
| Married | 73 | 76 | 84 | 86 |
| Widower | 15 | 12 | 7 | 3 |
| Divorced | 6 | 7 | 5 | 4 |
| Weight (kg) | 83 (75-92) | 84 (77-94) | 89 (81-98) | 92 (86-99) |
| Height (cm) | 159 (153-167) | 162 (155-170) | 168 (162-172) | 170 (165-174) |
| Body mass index (kg/m<sup>2</sup>) | 31.8 (29.8-34.4) | 31.6 (29.7-34.3) | 31.5 (29.6-33.4) | 32.3 (30.0-33.8) |
| Systolic blood pressure (mmHg) | 140 (130-153) | 138 (128-150) | 140 (129-150) | 140 (134-152) |
| Diastolic blood pressure (mmHg) | 77 (71-83) | 78 (71-85) | 80 (75-88) | 82 (77-91) |
| Smoking, % |  |  |  |  |
| Currently smoking | 5 | 11 | 10 | 13 |
| Former smoker (0-5 years ago) | 4 | 5 | 7 | 9 |
| Former smoker (>5 years ago) | 33 | 41 | 55 | 59 |
| Never smoker | 58 | 43 | 28 | 19 |
| Physical activity (METs-minutes/week) | 1692 (839-3224) | 1888 (839-3217) | 2238 (1259-3580) | 2098 (783-3238) |
| Diabetes, % | 43 | 28 | 19 | 20 |
| BECK depression inventory | 7 (4-13) | 7 (3-11) | 5 (3-12) | 5 (2-10) |
| Mediterranean diet adherence* | 8 (7-10) | 8 (6-10) | 6 (5-9) | 6 (4-7) |
| Daily wine consumption (drink/day) | 0 | 1 (1-1) | 2 (1-2) | 3 (2-3) |
| Daily beer consumption (drink/day) | 0 | 1 (0-1) | 1 (1-1) | 1 (0-1) |
| Daily spirits consumption (drink-day) | 0 | 0 (0-1) | 1 (0-1) | 1 (0-1) |
| *Biomarkers at baseline* |  |  |  |  |
| NT-proBNP | 57 (29-99) | 51 (26-92) | 47 (26-86) | 41 (25-78) |
| hsTnT | 7.6 (6.1-10.9) | 7.5 (6.0-10.7) | 9.2 (7.2-11.1) | 9.5 (7.6-11.9) |
| hsCRP | 0.3 (0.1-0.5) | 0.2 (0.1-0.4) | 0.2 (0.1-0.4) | 0.2 (0.1-0.4) |
| PICP | 614 (360-990) | 645 (362-1016) | 544 (332-978) | 496 (265-838) |
| 3-NT | 88 (68-110) | 87 (72-110) | 90 (78-113) | 89 (68-113) |

Presented as either median (25<sup>th</sup> percentile – 75<sup>th</sup> percentile) for continuous measures or as frequencies for categorical variables.
MET, metabolic equivalents of task. NT-proBNP, N-terminal pro-B-type natriuretic protein. hsTnT, high sensitivity troponin-T. hsCRP, high-sensitivity C-reactive protein. 3-NT, 3-nitrotyrosine. PICP, procollagen type 1 carboxy-terminal propeptide.
\*Sum of 16 questions concerning adherence to the energy-reduced Mediterranean diet. Does not include alcohol intake.
1 drink: 1-14 grams
2-3 drinks: 15-42 grams
4 or more drinks: 43 grams and more

Examination of cross-sectional associations of alcohol consumption with log-transformed biomarkers showed higher, but non-significant concentrations of PICP in those with higher alcohol consumption (β: 0.12, 95%CI: -0.02, 0.27) (**Table 2**). Overall, there were no associations present between baseline alcohol consumption and other baseline biomarkers.

**Table 2.** Association of baseline alcohol consumption with concentrations of log-transformed biomarkers at baseline, PREDIMED-Plus study.

|  |  | **0 drinks** | **1 drink/day** | **2-3 drinks/day** | **≥ 4 drinks/day** | **Per 1-drink difference** |
| --- | --- | --- | --- | --- | --- | --- |
| NT-proBNP | Model 1 | 0 (ref) | -0.01 (-0.20, 0.19) | 0.06 (-0.18, 0.30) | -0.02 (-0.30, 0.25) | -0.02 (-0.07, 0.04) |
|  | Model 2 | 0 (ref) | 0.02 (-0.17, 0.22) | 0.11 (-0.13, 0.35) | 0.03 (-0.25, 0.31) | -0.01 (-0.06, 0.05) |
| hsTnT | Model 1 | 0 (ref) | -0.04 (-0.13, 0.06) | 0.00 (-0.12, 0.11) | 0.04 (-0.09, 0.17) | 0.02 (-0.01, 0.04) |
|  | Model 2 | 0 (ref) | -0.02 (-0.11, 0.08) | 0.04 (-0.08, 0.15) | 0.07 (-0.07, 0.20) | 0.02 (0.00, 0.05) |
| hsCRP | Model 1 | 0 (ref) | -0.03 (-0.26, 0.19) | -0.01 (-0.28, 0.27) | -0.02 (-0.34, 0.30) | 0.00 (-0.06, 0.06) |
|  | Model 2 | 0 (ref) | -0.07 (-0.29, 0.15) | -0.03 (-0.31, 0.25) | -0.07 (-0.39, 0.25) | -0.01 (-0.07, 0.05) |
| PICP | Model 1 | 0 (ref) | 0.06 (-0.05, 0.16) | 0.13 (0.01, 0.26) | 0.12 (-0.02, 0.27) | 0.02 (-0.01, 0.05) |
|  | Model 2 | 0 (ref) | 0.05 (-0.05, 0.16) | 0.13 (0.00, 0.26) | 0.12 (-0.03, 0.26) | 0.02 (-0.01, 0.05) |
| 3-NT | Model 1 | 0 (ref) | 0.02 (-0.19, 0.23) | -0.02 (-0.28, 0.24) | -0.11 (-0.41, 0.19) | -0.03 (-0.09, 0.03) |
|  | Model 2 | 0 (ref) | 0.00 (-0.22, 0.21) | -0.06 (-0.33, 0.21) | -0.09 (-0.41, 0.21) | -0.03 (-0.09, 0.03) |

NT-proBNP, N-terminal pro-B-type natriuretic protein. hsTnT, high sensitivity troponin-T. high-sensitivity CRP, C-reactive protein. PICP, procollagen type 1 carboxy-terminal propeptide.
Model 1: multiple linear regression adjusted for age, sex, education.
Model 2: multiple linear regression adjusted for age, sex, education, marital status, smoking, physical activity, height, body mass index, systolic and diastolic blood pressure, diabetes, depressive symptoms, diet adherence.

Longitudinally, higher baseline alcohol consumption was associated with smaller increases in hsTnT concentration (β for 5-year change: -0.11, 95%CI: -0.20, -0.01 comparing ≥4 drinks/day to non-drinkers) (**Table 3 and Supplemental figure 1**). There was also evidence of (non-significant) smaller increases in PICP (β for 5-year change: -0.15, 95%CI: -0.30, 0.01) when comparing heavy-drinkers to non-consumers over the 5-year period. Associations showed similar patterns when visit was modeled as a categorical variable in the mixed models (**Supplemental Table 1**).

**Table 3.** Association of baseline alcohol consumption with 5-year change in log-transformed biomarkers from baseline to year 5, PREDIMED-Plus study.

|  |  | **0 drinks** | **1 drink/day** | **2-3 drinks/day** | **≥ 4 drinks/day** | **Per 1-drink difference** |
| --- | --- | --- | --- | --- | --- | --- |
| Change in total NT-proBNP | Model 1 | 0 (ref.) | 0.02 (-0.16, 0.21) | 0.08 (-0.14, 0.31) | 0.11 (-0.14, 0.37) | 0.04 (-0.01, 0.09) |
|  | Model 2 | 0 (ref.) | 0.03 (-0.16, 0.22) | 0.07 (-0.17, 0.30) | 0.08 (-0.19, 0.35) | 0.03 (-0.02, 0.08) |
| Change in hsTnT | Model 1 | 0 (ref.) | -0.05 (-0.11, 0.02) | -0.12 (-0.21, -0.04) | -0.12 (-0.21, -0.03) | -0.02 (-0.04, -0.01) |
|  | Model 2 | 0 (ref.) | -0.04 (-0.11, 0.03) | -0.11 (-0.19, -0.02) | -0.11 (-0.20, -0.01) | -0.02 (-0.04, 0.00) |
| Change in hsCRP | Model 1 | 0 (ref.) | 0.14 (-0.08, 0.35) | 0.17 (-0.09, 0.43) | 0.07 (-0.23, 0.37) | -0.01 (-0.06, 0.05) |
|  | Model 2 | 0 (ref.) | 0.16 (-0.07, 0.38) | 0.15 (-0.12, 0.43) | 0.06 (-0.26, 0.37) | -0.01 (-0.08, 0.05) |
| Change in PICP | Model 1 | 0 (ref.) | 0.08 (-0.03, 0.18) | 0.00 (-0.14, 0.13) | -0.12 (-0.27, 0.04) | -0.03 (-0.06, 0.00) |
|  | Model 2 | 0 (ref.) | 0.07 (-0.04, 0.18) | -0.03 (-0.17, 0.11) | -0.15 (-0.30, 0.01) | -0.04 (-0.07, -0.01) |
| Change in 3-NT | Model 1 | 0 (ref.) | 0.07 (-0.10, 0.23) | -0.08 (-0.28, 0.12) | 0.14 (-0.09, 0.37) | 0.01 (-0.03, 0.06) |
|  | Model 2 | 0 (ref.) | 0.07 (-0.10, 0.24) | -0.09 (-0.30, 0.12) | 0.14 (-0.10, 0.38) | 0.01 (-0.04, 0.06) |

NT-proBNP, N-terminal pro-B-type natriuretic protein. hsTnT, high sensitivity troponin-T. hsCRP, high-sensitivity C-reactive protein. 3-NT, 3-nitrotyrosine. PICP, procollagen type 1 carboxy-terminal propeptide.
Model 1: mixed linear model adjusted for age, sex, education, intervention group, and interaction of all covariates with time. Time is modeled in 5-year increments.
Model 2: mixed linear model adjusted for age, sex, education, intervention group, marital status, smoking, physical activity, height, body mass index, systolic and diastolic blood pressure, diabetes, depression, diet adherence, and interaction of all covariates with time. Time is modeled in 5-year increments.

In an analysis comparing those who increased their drinking amount to those who stayed the same over time, increasing alcohol consumption was associated with increases in hsCRP (β: 0.42, 95%CI: 0.11, 0.73), but not with other biomarkers (**Figure 2 and Table 4**).

**Table 4.** Association of changes in alcohol consumption with change in circulating log-transformed biomarkers from baseline to year 5, PREDIMED-Plus study.

|  |  | **Decreased consumption**<br>**(≥ 1 drinks/day)**<br>**(N = 126)** | **No change**<br>**(-1 > drinks/day > 1)**<br>**(N = 359)** | **Increased consumption**<br>**(≥ 1 drinks/day)**<br>**(N = 38)** | **Per 1-drink\* increase** |
| --- | --- | --- | --- | --- | --- |
| Change in total NT-proBNP | Model 1 | -0.01 (-0.23, 0.20) | 0 (ref.) | -0.15 (-0.42, 0.11) | 0.04 (-0.01, 0.09) |
|  | Model 2 | 0.00 (-0.22, 0.21) | 0 (ref.) | -0.14 (-0.41, 0.13) | 0.03 (-0.02, 0.08) |
| Change in hsTnT | Model 1 | -0.06 (-0.14, 0.01) | 0 (ref.) | -0.06 (-0.16, 0.03) | -0.02 (-0.04, 0.00) |
|  | Model 2 | -0.06 (-0.14, 0.01) | 0 (ref.) | -0.05 (-0.14, 0.05) | -0.02 (-0.04, 0.00) |
| Change in hsCRP | Model 1 | 0.09 (-0.15, 0.34) | 0 (ref.) | 0.41 (0.10, 0.71) | -0.01 (-0.06, 0.05) |
|  | Model 2 | 0.09 (-0.16, 0.34) | 0 (ref.) | 0.42 (0.11, 0.73) | -0.01 (-0.08, 0.05) |
| Change in PICP | Model 1 | 0.03 (-0.09, 0.16) | 0 (ref.) | -0.09 (-0.25, 0.06) | -0.03 (-0.06, 0.00) |
|  | Model 2 | 0.04 (-0.09, 0.16) | 0 (ref.) | -0.10 (-0.26, 0.05) | -0.04 (-0.07, -0.01) |
| Change in 3-NT | Model 1 | 0.04 (-0.16, 0.23) | 0 (ref.) | 0.03 (-0.20, 0.27) | 0.01 (-0.03, 0.06) |
|  | Model 2 | 0.03 (-0.16, 0.23) | 0 (ref.) | 0.02 (-0.22, 0.26) | 0.01 (-0.04, 0.06) |

\*1 drink = 14 grams of alcohol
NT-proBNP, N-terminal pro-B-type natriuretic protein. TnT, troponin-T. hsCRP, high-sensitivity C-reactive protein. 3-NT, 3-nitrotyrosine. PICP, procollagen type 1 carboxy-terminal propeptide.
Model 1: mixed linear model adjusted for age, sex, education, intervention group, and interaction of all covariates with time. Time is modeled in 5-year increments.
Model 2: mixed linear model adjusted for age, sex, education, intervention group, marital status, smoking, physical activity, height, body mass index, systolic and diastolic blood pressure, diabetes, depression, diet adherence, and interaction of all covariates with time. Time is modeled in 5-year increments.

**Figure 2.**
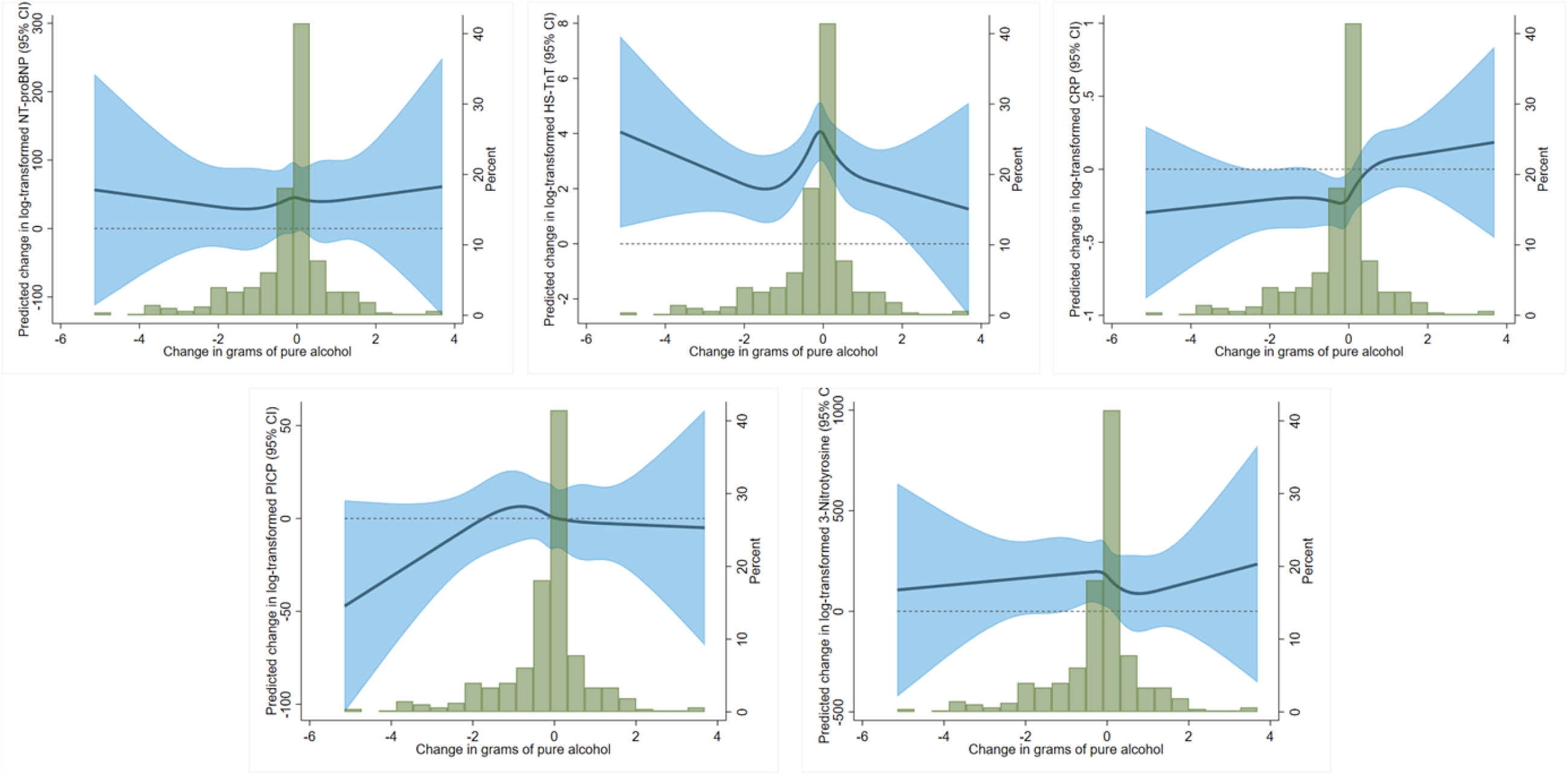
Associations of change in alcohol over 5 years modeled as restricted cubic splines with change in biomarkers, using the median change in alcohol consumption as the reference point. Top left to right: NT-proBNP, Hs-TnT, CRP. Bottom left to right: PICP, 3-NT. Multiple linear regression models adjusted for age, sex, education, and intervention group.

There was no evidence of differences between men and women in the baseline associations of alcohol consumption and cardiac biomarkers (**Supplemental table 2**). When looking at change in biomarkers over time, heavy drinking was associated with decreased hsTnT (β: -0.15, 95%CI: -0.28, -0.02) and increased (though not significantly) 3-NT (β: 0.33, 95%CI: 0.00, 0.67) in men, but not in women (**Supplemental table 3**). Once again, there was no evidence of statistically significant sex interactions.

## DISCUSSION

In this population at high-risk of cardiovascular disease, baseline alcohol consumption was not associated with cardiac biomarkers cross-sectionally. Heavy drinking at baseline, however, was associated with smaller increases in hsTnT and PICP by year 5. When accounting for changes in drinking behavior over 5 years, those who increased their alcohol consumption showed signs of increased inflammation, as evidenced by increased hsCRP levels.

PICP is an indicator of collagen production.^15^ The overproduction of collagen may induce cardiac fibrosis – the accumulation of proteins in the myocardial tissue.^16^ Severe fibrosis may leave the heart particularly vulnerable to cardiac remodeling. The balance between collagen synthesis and degradation may be impaired in AF and contribute to fibrogenesis in these patients.^17^ Furthermore, fibrotic damage to the heart – as indicated by elevated PICP – may precede left ventricular hypertrophy.^18^ Drinking more at baseline in this study showed signs of more fibrosis compared to low and moderate drinkers at baseline and over time, though not at statistically significant levels. Though we did not assess associations according to the type of drinks consumed in this analysis, wine – red, in particular – was the preferred drink in this Spanish cohort and promoted as part of the Mediterranean diet intervention. Resveratrol, (3,5,4′-trihydroxy-trans-stilbene), the most important type of stilbene, is a polyphenol highly present in red wine, and may play a role in mitigating myocardial fibrosis,^19-21^ potentially explaining the declining PICP levels in heavy drinkers over time. The needed amount of resveratrol to produce biologically relevant effects is probably very high and it cannot be obtained from the usual amounts contained in red wine, however. The amount from drinking wine is probably too low as to exert substantial cardioprotective effects and resveratrol effects are further hampered by its low bioavailability.^22^ However, resveratrol does not act in isolation. Other phenolic compounds, including other varieties of stilbenes, are also present in red wine may act synergistically, because they also exert similar bioactive beneficial effects. Among others, phenolic acids originated in the grapes pulp, anthocyanins and stilbenoids from the skin, and other phenols (catechins, proanthocyanidins and flavonols) usually present the skin and the seeds of grapes may contribute to these effects.

HsCRP is an indicator of overall inflammation in the body and is often a reliable gauge of cardiovascular health.^23,24^ General consensus on the link between alcohol consumption and inflammation, however, remains inconsistent, with some studies finding light-to-moderate drinkers having lower levels of inflammation compared to nondrinkers^25^ to U-shaped trends driven by former drinkers having higher levels of inflammation than their drinking counterparts.^26^ This inconsistency extends to its predictive value for AF.^27^

Oxidative and nitrosative stress are of particular concern in those that abuse alcohol.^28^ Protein nitration is the first step in a cellular cascade that can ultimately lead to cell death.^29^ The nitration of tyrosine (3-NT) is one way to induce damage in a cell and effectively stops the formation of critical enzymes and proteins.^16^ Atrial contractile dysfunction often seen in AF may in part be due to nitrative stress.^30^ Elevated levels of 3-NT were present in mice with alcohol-induced cardiomyopathy;^31^ many of these studies were done in vitro or in animals, less so in humans. In contrast, heavy drinking in this study was not associated with 3-NT at baseline nor with changes in 3-NT over time in this cohort of high-risk participants.

Moderate-to-heavy drinkers experienced smaller increases in hsTnT compared to abstainers over the 5-year follow-up period. Changes in drinking behavior did not appear to impact these findings. Our results seem to line up with findings from other cohorts, including ARIC, where moderate drinkers were less likely to experience incident increases in hsTnT compared to never drinkers;^32^ researchers found a similar effect in former drinkers compared to never drinkers.

PREDIMED-Plus did not collect data on drinking behavior before the start of the trial and heavy drinkers (>6 drinks/day) were excluded from the study, thus we cannot readily replicate findings in former drinkers.

Though no such associations were observed in the present study, NT-proBNP is an important indicator of cardiovascular health in the context of alcohol consumption.^33^ Moderate drinking was associated with elevated N-terminal pro-B-type natriuretic protein (NT-proBNP) in ARIC, cross-sectionally and prospectively over 6 years.^32^ Increased NT-proBNP is also associated with left ventricular hypertrophy (LVH) and ischemic events, though only in heavy drinkers.^34^ Chronically elevated NT-proBNP may increase systolic blood pressure and is a powerful biomarker of stroke risk.^35^ NT-proBNP is usually elevated in AF and is often a predictor of future AF.^36^ In a previous analysis of the PREDIMED-Plus trial, we found higher alcohol intake to be associated with larger left atrial volume and worsening atrial function, which may likely affect NT-proBNP concentrations.^37^ The apparent inconsistency between analyses evaluating circulating and imaging biomarkers of left atrial function supports the value of exploring pathophysiological pathways using multiple modalities.

Strengths of this study include having a well-characterized cohort nested in a large trial with excellent retention and alcohol and biomarker data available from multiple visits over the course of 5 years, and the availability of information on multiple potential confounders. We must also, however, address some of the limitations of this study. First, PREDIMED-Plus did not collect information on past drinking behavior, which may be biasing our current estimates. Second, though the FFQs have been previously validated, they are still dependent on participant recall, which may introduce some degree of misclassification in our study. Finally, generalizability of these findings to other populations, for example those at lower cardiovascular risk, is uncertain.

## CONCLUSION

In this population of participants at high risk of cardiovascular disease, alcohol consumption was associated with alterations in circulating biomarkers related to cardiovascular health. These findings contribute to a better understanding of the complex relationship between alcohol consumption and cardiovascular risk. However, further research is warranted to elucidate the precise molecular mechanisms, to determine the clinical implications of these observed changes in biomarker concentrations and to potentially inform public health guidelines on alcohol consumption, emphasizing the need for continued research in this area.

## Data Availability

Interested researchers can make a request for access to PREDIMED-Plus data directly from the study.

https://www.predimedplus.com/en/project/

## REFERENCES

1. Glantz MD, Bharat C, Degenhardt L, et al. The epidemiology of alcohol use disorders cross-nationally: Findings from the World Mental Health Surveys. Addict Behav 2020;102:106128.

2. Ronksley PE, Brien SE, Turner BJ, Mukamal KJ, Ghali WA. Association of alcohol consumption with selected cardiovascular disease outcomes: a systematic review and meta-analysis. BMJ 2011;342:d671.

3. Bell S, Daskalopoulou M, Rapsomaniki E, et al. Association between clinically recorded alcohol consumption and initial presentation of 12 cardiovascular diseases: population based cohort study using linked health records. BMJ 2017;356:j909.

4. Groh CA, Faulkner M, Getabecha S, et al. Patient-reported triggers of paroxysmal atrial fibrillation. Heart Rhythm 2019;16:996–1002.

5. Voskoboinik A, Kalman JM, De Silva A, et al. Alcohol Abstinence in Drinkers with Atrial Fibrillation. N Engl J Med 2020;382:20–8.

6. Zagrosek A, Messroghli D, Schulz O, Dietz R, Schulz-Menger J. Effect of binge drinking on the heart as assessed by cardiac magnetic resonance imaging. JAMA 2010;304:1328–30.

7. Voskoboinik A, Wong G, Lee G, et al. Moderate alcohol consumption is associated with atrial electrical and structural changes: Insights from high-density left atrial electroanatomic mapping. Heart Rhythm 2019;16:251–9.

8. Voskoboinik A, Costello BT, Kalman E, et al. Regular Alcohol Consumption Is Associated With Impaired Atrial Mechanical Function in the Atrial Fibrillation Population: A Cross-Sectional MRI-Based Study. JACC Clin Electrophysiol 2018;4:1451–9.

9. Di Castelnuovo A, Costanzo S, Bagnardi V, Donati MB, Iacoviello L, de Gaetano G. Alcohol dosing and total mortality in men and women: an updated meta-analysis of 34 prospective studies. Arch Intern Med 2006;166:2437–45.

10. Di Castelnuovo A, Costanzo S, Bonaccio M, et al. Moderate Alcohol Consumption Is Associated With Lower Risk for Heart Failure But Not Atrial Fibrillation. JACC Heart Fail 2017;5:837–44.

11. Martinez-Gonzalez MA, Buil-Cosiales P, Corella D, et al. Cohort Profile: Design and methods of the PREDIMED-Plus randomized trial. Int J Epidemiol 2019;48:387–8o.

12. Fernandez-Ballart JD, Pinol JL, Zazpe I, et al. Relative validity of a semi-quantitative food-frequency questionnaire in an elderly Mediterranean population of Spain. Br J Nutr 2010;103:1808–16.

13. Schroder H, Cardenas-Fuentes G, Martinez-Gonzalez MA, et al. Effectiveness of the physical activity intervention program in the PREDIMED-Plus study: a randomized controlled trial. Int J Behav Nutr Phys Act 2018;15:110.

14. Schroder H, Zomeno MD, Martinez-Gonzalez MA, et al. Validity of the energy-restricted Mediterranean Diet Adherence Screener. Clin Nutr 2021;40:4971–9.

15. Fan D, Takawale A, Lee J, Kassiri Z. Cardiac fibroblasts, fibrosis and extracellular matrix remodeling in heart disease. Fibrogenesis Tissue Repair 2012;5:15.

16. Deng XS, Deitrich RA. Ethanol metabolism and effects: nitric oxide and its interaction. Curr Clin Pharmacol 2007;2:145–53.

17. Lofsjogard J, Persson H, Diez J, et al. Atrial fibrillation and biomarkers of myocardial fibrosis in heart failure. Scand Cardiovasc J 2014;48:299–303.

18. Ho CY, Lopez B, Coelho-Filho OR, et al. Myocardial fibrosis as an early manifestation of hypertrophic cardiomyopathy. N Engl J Med 2010;363:552–63.

19. Feng H, Mou SQ, Li WJ, et al. Resveratrol Inhibits Ischemia-Induced Myocardial Senescence Signals and NLRP3 Inflammasome Activation. Oxid Med Cell Longev 2020;2020:2647807.

20. Liu J, Zhang M, Qin C, et al. Resveratrol Attenuate Myocardial Injury by Inhibiting Ferroptosis Via Inducing KAT5/GPX4 in Myocardial Infarction. Front Pharmacol 2022;13:906073.

21. Jiang J, Gu X, Wang H, Ding S. Resveratrol improves cardiac function and left ventricular fibrosis after myocardial infarction in rats by inhibiting NLRP3 inflammasome activity and the TGF-beta1/SMAD2 signaling pathway. PeerJ 2021;9:e11501.

22. Tome-Carneiro J, Gonzalvez M, Larrosa M, et al. Resveratrol in primary and secondary prevention of cardiovascular disease: a dietary and clinical perspective. Ann N Y Acad Sci 2013;1290:37–51.

23. Albert CM, Ma J, Rifai N, Stampfer MJ, Ridker PM. Prospective study of C-reactive protein, homocysteine, and plasma lipid levels as predictors of sudden cardiac death. Circulation 2002;105:2595–9.

24. Ridker PM, Rifai N, Rose L, Buring JE, Cook NR. Comparison of C-reactive protein and low-density lipoprotein cholesterol levels in the prediction of first cardiovascular events. N Engl J Med 2002;347:1557–65.

25. Iakunchykova O, Averina M, Kudryavtsev AV, et al. Evidence for a Direct Harmful Effect of Alcohol on Myocardial Health: A Large Cross-Sectional Study of Consumption Patterns and Cardiovascular Disease Risk Biomarkers From Northwest Russia, 2015 to 2017. J Am Heart Assoc 2020;9:e014491.

26. Averina M, Nilssen O, Arkhipovsky VL, Kalinin AG, Brox J. C-reactive protein and alcohol consumption: Is there a U-shaped association? Results from a population-based study in Russia. The Arkhangelsk study. Atherosclerosis 2006;188:309–15.

27. Sinner MF, Stepas KA, Moser CB, et al. B-type natriuretic peptide and C-reactive protein in the prediction of atrial fibrillation risk: the CHARGE-AF Consortium of community-based cohort studies. Europace 2014;16:1426–33.

28. Di Luzio NR. A mechanism of the acute ethanol-induced fatty liver and the modification of liver injury by antioxidants. Am J Pharm Sci Support Public Health 1966;15:50–63.

29. Sultana R, Ravagna A, Mohmmad-Abdul H, Calabrese V, Butterfield DA. Ferulic acid ethyl ester protects neurons against amyloid beta-peptide(1-42)-induced oxidative stress and neurotoxicity: relationship to antioxidant activity. J Neurochem 2005;92:749–58.

30. Mihm MJ, Yu F, Carnes CA, et al. Impaired myofibrillar energetics and oxidative injury during human atrial fibrillation. Circulation 2001;104:174–80.

31. Tan Y, Li X, Prabhu SD, et al. Angiotensin II plays a critical role in alcohol-induced cardiac nitrative damage, cell death, remodeling, and cardiomyopathy in a protein kinase C/nicotinamide adenine dinucleotide phosphate oxidase-dependent manner. J Am Coll Cardiol 2012;59:1477–86.

32. Lazo M, Chen Y, McEvoy JW, et al. Alcohol Consumption and Cardiac Biomarkers: The Atherosclerosis Risk in Communities (ARIC) Study. Clin Chem 2016;62:1202–10.

33. Britton A, O’Neill D, Kuh D, Bell S. Sustained heavy drinking over 25 years is associated with increased N-terminal-pro-B-type natriuretic peptides in early old age: Population-based cohort study. Drug Alcohol Depend 2020;212:108048.

34. Wentzel A, Malan L, Scheepers JD, Malan NT. QTc prolongation, increased NT-proBNP and pre-clinical myocardial wall remodeling in excessive alcohol consumers: The SABPA study. Alcohol 2018;68:1–8.

35. Collaboration NPS, Willeit P, Kaptoge S, et al. Natriuretic peptides and integrated risk assessment for cardiovascular disease: an individual-participant-data meta-analysis. Lancet Diabetes Endocrinol 2016;4:840–9.

36. Patton KK, Ellinor PT, Heckbert SR, et al. N-terminal pro-B-type natriuretic peptide is a major predictor of the development of atrial fibrillation: the Cardiovascular Health Study. Circulation 2009;120:1768–74.

37. Alam AB, Toledo E., Romaguera D., Alonso-Gómez A.M., Martínez-González M.A., Tojal-Sierra L., Noris Mora M., Mas-Llado C., Li L., Gonzalez Casanova I., Salas-Salvadó J., Fitó M., Alonso A. Associations of alcohol consumption with left atrial morphology and function in a population at high cardiovascular risk. medRxiv 2023.

