## Supplemental Tables for "Association of alcohol consumption with circulating biomarkers of atrial fibrillation-related pathways in a population at high cardiometabolic risk"

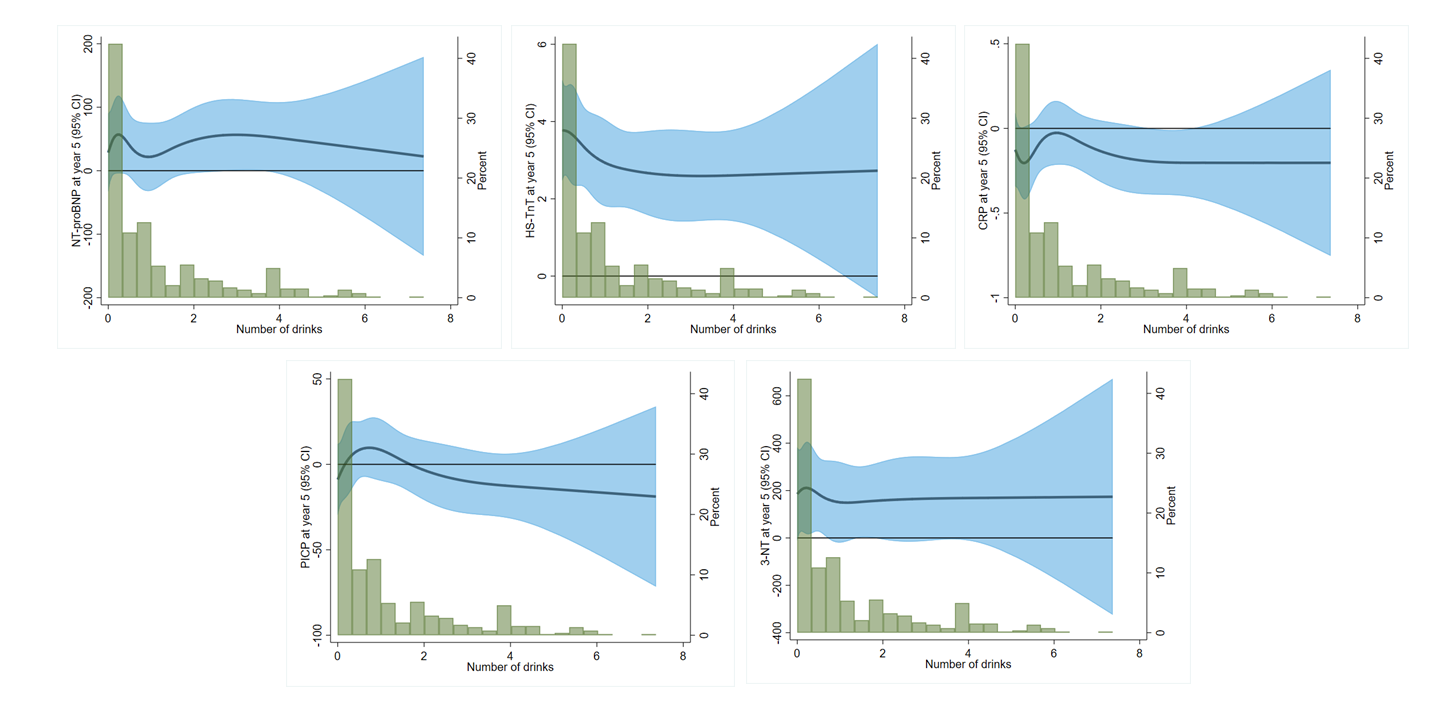

Supplemental Figure 1. Associations of baseline alcohol consumption modeled as restricted cubic splines with change in biomarkers. Top left to right: NT-proBNP, Hs-TnT, CRP. Bottom left to right: PICP, 3-NT. Multiple lineaer regression models adjusted for age, sex, education, and intervention group.

Supplemental Table 1. Association of baseline alcohol consumption (per 1 drink/day difference) with change in log-transformed biomarkers, PREDIMED-Plus

|  | Per 1 drink/day difference | |
| --- | --- | --- |
|  | Year 3 vs baseline | Year 5 vs baseline |
| NT-proBNP | 0.07 (0.01, 0.12) | 0.02 (-0.03, 0.07) |
| hsTnT | 0.00 (-0.02, 0.02) | -0.02 (-0.04, 0.00) |
| hsCRP | -0.03 (-0.09, 0.03) | -0.01 (-0.07, 0.05) |
| PICP | -0.02 (-0.05, 0.01) | -0.04 (-0.07, -0.01) |
| 3-NT | 0.00 (-0.05, 0.05) | 0.01 (-0.03, 0.06) |

NT-proBNP, N-terminal pro-B-type natriuretic protein. TnT, troponin-T. hsCRP, high-sensitivity C-reactive protein. PICP, procollagen type 1 carboxy-terminal propeptide

Mixed linear model adjusted for age, sex, education, intervention group, marital status, smoking, physical activity, height, body mass index (BMI), systolic and diastolic blood pressure, diabetes, depression, diet adherence, and interaction of all covariates with time. Time modeled as a categorical variable (baseline, year 3, year 5).

Supplemental Table 2. Multiple linear regression estimates of overall alcohol consumption with log-transformed biomarkers at baseline, by sex, PREDIMED-Plus.

|  | **0 drinks** | **1 drink/day** | **2 drinks/day** | **≥ 3 drinks/day** | **Per 1-drink difference** |
| --- | --- | --- | --- | --- | --- |
| **Male = 272** | | | | | |
| NT-proBNP | -0.17 (-0.50, 0.15) | -0.17 (-0.50, 0.15) | -0.17 (-0.50, 0.15) | -0.17 (-0.50, 0.15) | -0.03 (-0.10, 0.04) |
| TnT | 0.11 (-0.04, 0.27) | 0.11 (-0.04, 0.27) | 0.11 (-0.04, 0.27) | 0.11 (-0.04, 0.27) | 0.02 (-0.01, 0.06) |
| hsCRP | 0.06 (-0.32, 0.45) | 0.06 (-0.32, 0.45) | 0.06 (-0.32, 0.45) | 0.06 (-0.32, 0.45) | -0.05 (-0.13, 0.02) |
| PICP | 0.14 (-0.04, 0.31) | 0.14 (-0.04, 0.31) | 0.14 (-0.04, 0.31) | 0.14 (-0.04, 0.31) | 0.04 (0.00, 0.07) |
| 3-nitrotyrosine | -0.35 (-0.72, 0.03) | -0.35 (-0.72, 0.03) | -0.35 (-0.72, 0.03) | -0.35 (-0.72, 0.03) | -0.06 (-0.14, 0.01) |
| **Female = 190** | | | | | |
| NT-proBNP | 0.11 (-0.15, 0.35) | 0.11 (-0.15, 0.35) | 0.11 (-0.15, 0.35) | 0.11 (-0.15, 0.35) | 0.08 (-0.13, 0.29) |
| TnT | -0.10 (-0.23, 0.02) | -0.10 (-0.23, 0.02) | -0.10 (-0.23, 0.02) | -0.10 (-0.23, 0.02) | 0.09 (-0.01, 0.19) |
| hsCRP | -0.09 (-0.34, 0.16) | -0.09 (-0.34, 0.16) | -0.09 (-0.34, 0.16) | -0.09 (-0.34, 0.16) | -0.08 (-0.27, 0.10) |
| PICP | 0.01 (-0.13, 0.15) | 0.01 (-0.13, 0.15) | 0.01 (-0.13, 0.15) | 0.01 (-0.13, 0.15) | 0.10 (-0.01, 0.22) |
| 3-nitrotyrosine | 0.17 (-0.09, 0.43) | 0.17 (-0.09, 0.43) | 0.17 (-0.09, 0.43) | 0.17 (-0.09, 0.43) | 0.00 (-0.20, 0.20) |
| *Sex interactions* | | | | | |
| NT-proBNP | | P = 0.26 | | | |
| TnT | | P = 0.64 | | | |
| CRP | | P = 0.58 | | | |
| PICP | | P = 0.46 | | | |
| 3-nitrotyrosine | | P = 0.63 | | | |

NT-proBNP, N-terminal pro-B-type natriuretic protein. TnT, troponin-T. hsCRP, high-sensitivity C-reactive protein. PICP, procollagen type 1 carboxy-terminal propeptide.

All estimates are from model 2. Model 2 adjusts for age, education, marital status, smoking, physical activity, height, body mass index (BMI), systolic and diastolic blood pressure, diabetes, depression, and diet adherence.

Supplemental Table 3. Multiple linear regression estimates of overall alcohol consumption at baseline with 5-year change in log-transformed biomarkers from baseline to year 5, by sex, PREDIMED-Plus.

|  | **0 drinks** | **1 drink/day** | **2 drinks/day** | **> 2 drinks/day** | **Per 1-drink difference** |
| --- | --- | --- | --- | --- | --- |
| **Male = 272** | | | | | |
| NT-proBNP | 0.06 (-0.24, 0.37) | 0.06 (-0.24, 0.37) | 0.06 (-0.24, 0.37) | 0.06 (-0.24, 0.37) | 0.04 (-0.02, 0.10) |
| TnT | -0.06 (-0.18, 0.06) | -0.06 (-0.18, 0.06) | -0.06 (-0.18, 0.06) | -0.06 (-0.18, 0.06) | -0.03 (-0.06, -0.01) |
| hsCRP | 0.10 (-0.32, 0.51) | 0.10 (-0.32, 0.51) | 0.10 (-0.32, 0.51) | 0.10 (-0.32, 0.51) | 0.00 (-0.09, 0.08) |
| PICP | 0.05 (-0.14, 0.23) | 0.05 (-0.14, 0.23) | 0.05 (-0.14, 0.23) | 0.05 (-0.14, 0.23) | -0.06 (-0.10, -0.02) |
| 3-nitrotyrosine | 0.21 (-0.09, 0.51) | 0.21 (-0.09, 0.51) | 0.21 (-0.09, 0.51) | 0.21 (-0.09, 0.51) | 0.06 (0.00, 0.12) |
| **Female = 190** | | | | | |
| NT-proBNP | 0.00 (-0.27, 0.26) | 0.00 (-0.27, 0.26) | 0.00 (-0.27, 0.26) | 0.00 (-0.27, 0.26) | 0.00 (-0.21, 0.21) |
| TnT | -0.02 (-0.10, 0.07) | -0.02 (-0.10, 0.07) | -0.02 (-0.10, 0.07) | -0.02 (-0.10, 0.07) | -0.06 (-0.13, 0.00) |
| hsCRP | 0.19 (-0.05, 0.44) | 0.19 (-0.05, 0.44) | 0.19 (-0.05, 0.44) | 0.19 (-0.05, 0.44) | 0.10 (-0.08, 0.28) |
| PICP | 0.09 (-0.07, 0.24) | 0.09 (-0.07, 0.24) | 0.09 (-0.07, 0.24) | 0.09 (-0.07, 0.24) | 0.00 (-0.13, 0.12) |
| 3-nitrotyrosine | 0.04 (-0.17, 0.25) | 0.04 (-0.17, 0.25) | 0.04 (-0.17, 0.25) | 0.04 (-0.17, 0.25) | -0.08 (-0.24, 0.08) |
| *Sex interactions* | | | | | |
| NT-proBNP | | P = 0.40 | | | |
| TnT | | P = 0.19 | | | |
| CRP | | P = 0.52 | | | |
| PICP | | P = 0.61 | | | |
| 3-nitrotyrosine | | P = 0.41 | | | |

NT-proBNP, N-terminal pro-B-type natriuretic protein. TnT, troponin-T. hsCRP, high-sensitivity C-reactive protein. PICP, procollagen type 1 carboxy-terminal propeptide.

All estimates are from model 2. Model 2 adjusts for age, education, marital status, smoking, physical activity, height, body mass index (BMI), systolic and diastolic blood pressure, diabetes, depression, diet adherence, and interaction of all covariates with time. Time is modeled in 5-year increments.
